# Foundation Models Reveal Untapped Health Information in Human Polysomnographic Sleep Data

**DOI:** 10.1101/2025.07.15.25331562

**Authors:** William G. Coon, Mattson Ogg

## Abstract

Traditional sleep assessment methods rely on visual scoring of polysomnography (PSG), categorizing sleep into discrete stages (Wake, N1, N2, N3, REM) based on standardized guidelines developed by Rechtschaffen and Kales and later refined by the American Academy of Sleep Medicine (AASM). While undeniably useful, these methods are limited by subjectivity, arbitrary thresholds, and coarse temporal resolution, potentially overlooking important physiological information embedded within sleep EEG data. Recent advances in artificial intelligence, particularly Foundation Models (FM) trained through self-supervised learning (SSL), provide promising opportunities to address these limitations by autonomously uncovering rich, data-driven representations of sleep structure. In this study, we applied a SSL approach to large-scale PSG datasets, enabling the identification of latent sleep states and transitional dynamics not fully represented by traditional scoring methods. We found that these SSL-derived representations substantially enhanced predictive accuracy for critical health metrics, including demographics, clinical sleep metrics, mood, and cognitive performance. Comparisons demonstrated that this predictive advantage was not merely due to neural network complexity, as models trained directly from scratch without SSL pretraining significantly underperformed, nor was it due to having internalized simply any representation of sleep, as models pretrained to internalize traditional sleep stages could not match the SSL-pretrained model’s predictive accuracy. Taken together, these results indicate that FMs can identify structure from sleep data that encodes more than traditional sleep stage structure, uncovering previously untapped health information. These findings underscore the considerable potential of Foundation Models and SSL techniques to extend, complement, and enrich traditional sleep analyses, offering deeper insights into sleep’s complex relationship with overall health and cognitive function.

## 1. Introduction

Sleep, traditionally assessed through visual scoring of polysomnography (PSG), has been categorized into discrete stages (most recently Wake, N1, N2, N3, REM) based on standards originally developed in the 1960s by Rechtschaffen and Kales and later refined by the American Academy of Sleep Medicine (AASM) in 2007 (Kales and Rechtschaffen 1968; Silber et al. 2007). These visual scoring guidelines have been instrumental in standardizing sleep research and remain a cornerstone of sleep medicine. They also come with long-recognized limitations –many of which were acknowledged first by the authors of the guidelines themselves– that constrain their clinical and scientific utility (Kales and Rechtschaffen 1968; Silber et al. 2007; Himanen and Hasan 2000; Schulz 2008; van Gorp et al. 2022). For example, reliance on human visual judgment introduces subjectivity, resulting in considerable variability among scorers (Danker-Hopfe et al. 2009; Rosenberg and Van Hout 2013). Fixed criteria such as the 75 µV amplitude threshold for defining slow-wave activity may not generalize well across diverse populations or accurately reflect physiological changes associated with aging (Davidson et al. 2025). Furthermore, the traditional emphasis on visually prominent signals is insensitive to subtle but potentially important physiological markers that may not be apparent to the naked eye. The fixed 30-second scoring epoch, a legacy of earlier recording technologies, limits the ability to capture shorter-lived physiological phenomena or micro-states, which could hold significant functional implications (Brodbeck et al. 2012). Lastly, traditional methods inadequately address continuous physiological transitions and the inherent variability within each sleep stage, making them less effective for conditions characterized by fragmented or atypical sleep patterns (Decat et al. 2022; Stephansen et al. 2018). These limitations persist even with contemporary automated scoring algorithms (Coon, Zerr, et al. 2025; Coon and Ogg 2024; Coon and Punjabi 2021; Coon, Luna, et al. 2025; Perslev et al. 2021; W. Zhang et al. 2024; Yao et al. 2023; Phan et al. 2022; M. Kim et al. 2023; D. Kim et al. 2022; Stephansen et al. 2018; Korkalainen et al. 2020; Hanna and Flöel 2023), which precisely learn human-defined criteria rather than revealing deeper physiological patterns discoverable by data-driven techniques (Huijben et al. 2025; 2023; van Gorp et al. 2022; Decat et al. 2022; Banville et al. 2021). In summary, traditional scoring guidelines have provided the foundation for clinical sleep medicine to date and will remain essential in clinical and research settings, yet inherently overlook neurophysiological information contained within sleep signals. This information will remain untapped until traditional sleep stages can be augmented by pairing them with complimentary state information.

Recent advances in artificial intelligence, particularly Foundation Models (FMs) and self-supervised learning (SSL), present significant opportunities to extend, enrich, and complement traditional sleep scoring methods (Fox et al. 2025; Ogg and Coon 2024; Ogg et al. 2025; Thapa et al. 2024). SSL techniques empower neural networks to autonomously identify intrinsic structural patterns within large-scale datasets (Krishnan et al. 2022). This process occurs without any reliance on data labels (e.g., sleep stage labels), allowing models to capture nuanced and complex sleep dynamics beyond visually defined features and without any of the biases or shortcomings of previously defined scoring criteria. By learning rich and generalizable representations of sleep’s latent physiological structure, SSL-trained FMs can uncover novel states and transition dynamics that traditional scoring frameworks overlook, thereby providing new insights into sleep’s complex relationship with overall health, cognitive function, and mental health.

In this report, we demonstrate that Foundation Models trained using SSL can meaningfully contribute to developing more comprehensive, data-driven schemas for sleep analysis and classification. Our approach identifies physiological heterogeneity within traditionally defined sleep stages that contains untapped health information, demonstrated by enhanced predictive capabilities for important health-related outcomes such as brain health, cognitive performance, and mood disorders. Crucially, these predictive strengths originate from the latent structure discovered through SSL (i.e., sleep structure) rather than from the architecture of the neural network itself, as evidenced by comparative analyses showing that identical model architectures trained without SSL-derived pre-training fail to achieve comparable outcomes. Our results demonstrate that Foundation Models can autonomously extract previously untapped health information. They also underscore the potential of leveraging SSL and large-scale datasets to refine our understanding of sleep physiology, ultimately providing deeper insights into its implications for human health and disease.

## 2. Methods

### 2.1. Data and Preprocessing

We compiled polysomnography (PSG) data from multiple large-scale publicly available datasets, totaling over 11,000 individual records, for use in training, validation, and testing. Data were sourced from the National Sleep Research Resource (NSRR) (G.-Q. Zhang et al. 2018), with the resulting training, validation, and testing partitions summarized in Table 1. Specifically, seven datasets (Lee et al. 2022; Chen et al. 2015; Redline et al. 2012; Blackwell et al. 2011; Young et al. 2009; Spira et al. 2008; Quan et al. 1997) were combined to form the model training set, which comprised 11,261 PSG records obtained from 7,201 individuals.

**Table 1.** Datasets and Demographics.

| Dataset | Age<br>(Mean/Mdn) | Age Range<br>(Min./Max.) | Sex (F/M) | Participants | Overnights | Total<br>Examples |
| --- | --- | --- | --- | --- | --- | --- |
| MESA | 69.42/68 | 54/90 | 991/861 | 1,852 | 1,852 | 87,366 |
| MrOS | 74.67/74 | 67/90 | 0/903 | 903 | 1,808 | 96,713 |
| SHHS | 62.47/62 | 39/90 | 1,229/1,071 | 2,300 | 4,698 | 191,880 |
| WSC | 58.16/58 | 37/78 | 438/481 | 919 | 1,660 | 53,989 |
| SOF | 83/82 | 76/90 | 393/0 | 398 | 398 | 17,581 |
| CFS | 41.44/43.31 | 6.77/88.53 | 362/295 | 657 | 657 | 28,826 |
| NCHSDB | 21.38/19.93 | 18/58.35 | 88/84 | 172 | 188 | 5,915 |
| Train Total | 63.15/65 | 6.77/90 | 3,506/3,695 | 7,201 | 11,261 | 482,270 |
| Held-Out Train | 63.37/65.5 | 10.76/90 | 381/398 | 779 | 779 | 32,405 |
| Dreem* | 35.32/- | -/- | 6/19 | 80 | 80 | 742 |
| Sleep-EDF | 59.21/57 | 25/101 | 41/36 | 77 | 152 | 1,849 |
| HomePAP | 46.05/45.98 | 20.25/79.77 | 92/99 | 191 | 191 | 1,668 |
| APPLES | 51.59/52 | 18/83 | 353/691 | 1,044 | 1,044 | 9,683 |

We held out an "internal" validation partition of records from 779 randomly selected participants from the seven datasets used for training (roughly 10% of the total participants, one record each). This internal validation set closely mirrored the demographic characteristics of the training dataset. All other datasets were held out from training and validation to assess generalization performance and for downstream experiments. This included smaller standard benchmark corpora like Dreem (Guillot et al. 2020) and Sleep-EDF (Kemp et al. 2000; Goldberger et al. 2000) for preliminary evaluation and exploration of model representations, although these do not include a full multimodal set of PSG signals used for some models (e.g., they lack respiratory and/or ECG signals). We also selected the in-laboratory HomePAP dataset (Rosen et al. 2012), as a hold out dataset for some experiments as it frequently serves as a benchmark for external evaluation in prior work (Brink-Kjaer et al. 2022; D. Zhang et al. 2024). For the final test dataset, we selected the publicly available APPLES dataset (Quan et al. 2011; Kushida et al. 2012), specifically the available data from Visit 3 ("DX"). This dataset was chosen due to its extensive baseline physiological data along with detailed cognitive and mood assessments. Cognitive and clinical assessments retained for analysis included: Profile of Mood States (POMS; McNair, Lorr, and Droppleman 1971), Epworth Sleepiness Scale (ESS; Johns 1991), Buschke Selective Reminding Test (BSRT; Buschke 1973), Pathfinder Task (Malanchini et al. 2021), Psychomotor Vigilance Task (PVT; Dinges and Powell 1985), and the midday Sustained Working Memory Test (SWMT; Gevins et al. 2011, all from Visit 3), as well as Wechsler Abbreviated Scale of Intelligence IQ (WASI-IQ; Wechsler 2012), Mini-Mental State Exam (MMSE; (Folstein, Folstein, and McHugh 1975), Beck Depression Inventory (BDI; Beck et al. 1961), Hamilton Rating Scale for Depression (HAM-D; Hamilton 1960), Body Mass Index (BMI), and the apnea-hypopnea index (AHI; all obtained during the baseline visit).

From each PSG recording, we extracted time series signals from a central EEG channel (C3 or C4), as well as EOG (left or right), EMG (from the chin), ECG, and the thoracic respiration band channels. These were each resampled to 100 Hz and robustly normalized (median-centered, scaled by the signal’s interquartile range, and clipped within ±20 times the IQR). Preprocessing steps were conducted using MNE-Python (Gramfort 2013).

Data were then segmented into 30-second epochs, retaining only epochs assigned a sleep stage label according to standardized scoring guidelines (e.g., AASM; Silber et al. 2007). This selective retention was performed to enhance data quality for comparative analyses. After segmentation, epochs were assembled into sequences of 101 consecutive epochs, representing 50.5 minutes of data per sequence, for model input. This duration was chosen to balance comprehensive exposure to typical sleep-cycle patterns and fine-grained temporal resolution of the input sequence. Training data sequences from the seven datasets were formed using a 25-epoch hop size to augment dataset size (approximately four-fold oversampling) and to expose the model to a variety of different “views” of captured sleep cycles during training. In contrast, sequences from the external Dreem, Sleep-EDF, HomePAP and APPLES datasets were generated without overlap for unbiased evaluation. Any PSG records or sequences with missing signals were excluded to maintain comparability across PSG signal modalities.

### 2.2. Self-Supervised Learning

We implemented a self-supervised learning (SSL) approach inspired by the HuBERT methodology (Hsu et al. 2021) developed for speech analysis, wherein a transformer model is trained to predict sequences of discrete k-means-derived labels obtained from the training dataset. A schematic representation of this approach is provided in Figure 1. To derive these labels, each 101-epoch EEG time-series segment was transformed into time-frequency power distributions using 30-second time bins computed using the YASA toolbox (Vallat and Walker 2021). This clustering solution was fit using a mini-batch k-means algorithm applied to the spectral information of each epoch from one randomly selected 101-epoch segment per participant from the training dataset (the k-means model was fit to a subset of the data due to the large memory footprint of the time-frequency decompositions, similar to the original approach; Hsu et al. 2021). The resulting cluster assignments were then systematically applied to all 30-second training and validation epochs based on their time-frequency content. Models that took additional signals as input (e.g., EEG+EOG in one model, or EEG+EOG+EMG+ECG+respiration in the “PSG” model) followed a similar procedure but time-frequency decompositions for all input signals were concatenated (in the frequency dimension) before input to the k-means model (i.e., producing a matrix with rows of spectral features from each signal for each 101-epoch time bin column).

**Fig. 1.**
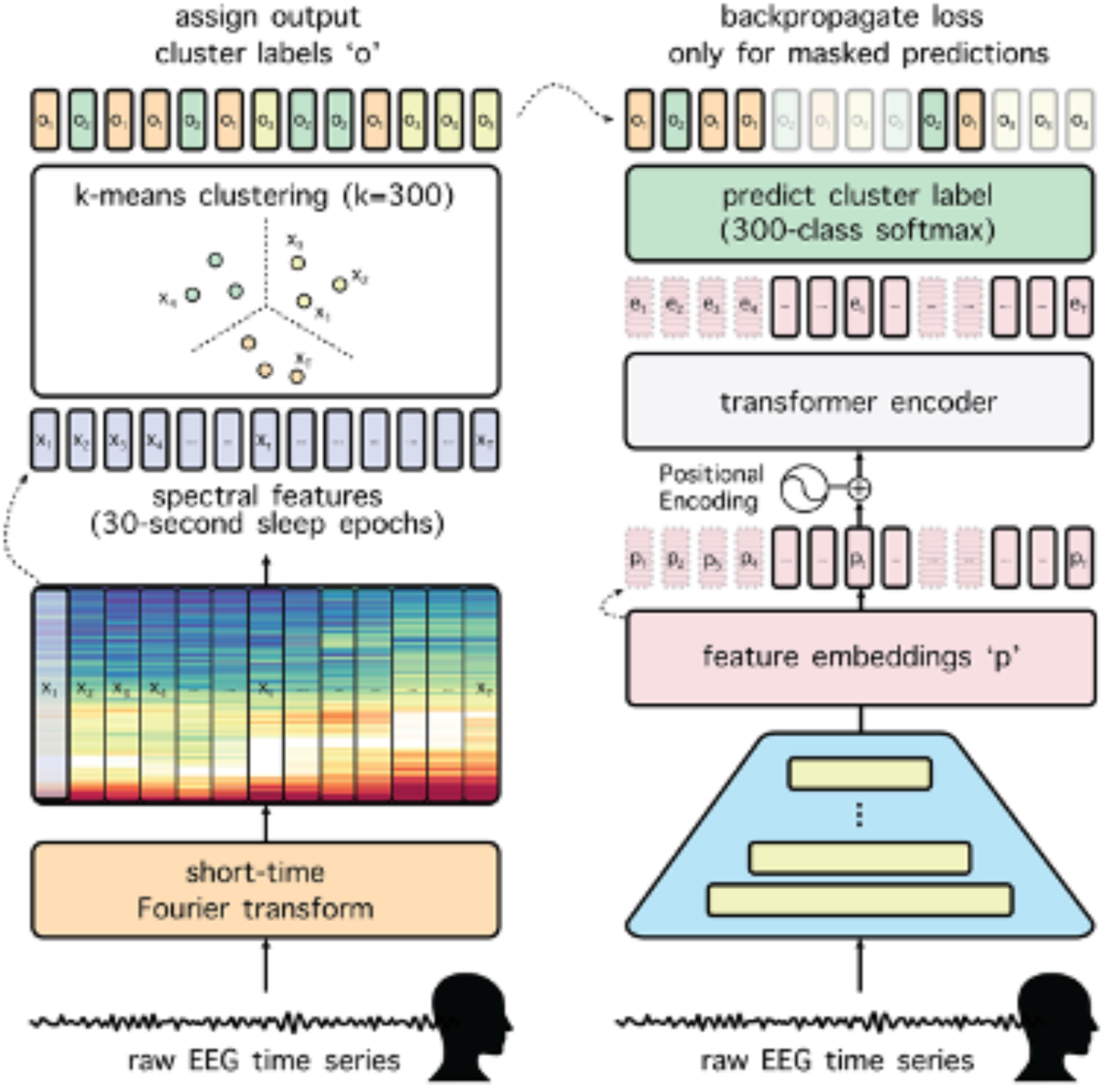
Self-supervised training approach. Models trained via self-supervised learning used an approach inspired by HuBERT (Hsu et al. 2021), in which SSL models were trained using pseudo-labels derived from k-means clustering of the time-frequency representations of 30-second sleep epochs. These pseudo-labels served as classification targets during pretraining. Similar to HuBERT, models predicted the pseudo-labels of randomly masked segments of the input signal using information from surrounding, unmasked portions. Masking encourages the model to learn robust and generalized features by preventing reliance on local signal continuity, thereby enhancing SSL model performance.

During the self-supervised training phase, portions of the input data corresponding to segments of the k-means label sequences were masked (set to zero) before processing by the transformer layers. Masking occurred in contiguous blocks of 10 epochs, with each epoch within the 101-epoch sequence having an 8% chance of initiating a masked block. The model was trained to predict the original k-means labels for these masked segments, minimizing cross-entropy loss.

The model architecture (Fig. 2 [model architecture]) began with a convolutional front end comprising series of seven one-dimensional convolutional layers, each containing 512 channels, with strides of [5, 2, 2, 2, 2, 2, 2] and kernel widths of [21, 3, 3, 3, 3, 2, 2]. Each convolutional layer was followed by layer normalization and GELU activation. The output from the convolutional layers was subsequently projected linearly from 512 to 768 dimensions, activated by GELU, and then passed through positional encoding. The positional-encoded data were then input to 12 transformer encoder layers, each equipped with 12 attention heads, an internal feed-forward dimension of 3,072, GELU activations, and a 5% layer dropout probability.

**Fig. 2.**
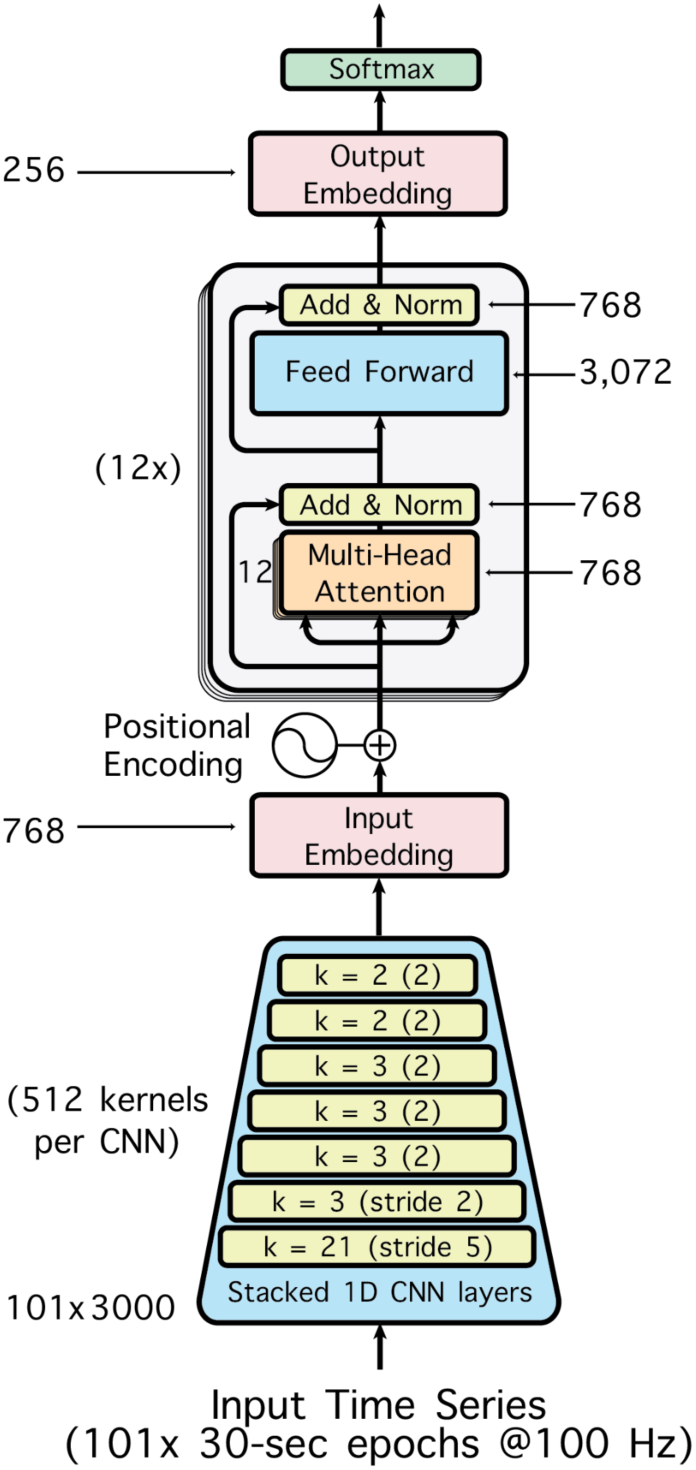
Model architecture. All models used the same underlying architecture. Signal features are extracted by a convolutional frontend, positionally encoded, and submitted to a transformer stack, before final submission to a softmax classification head to predict either sleep stage label or cluster pseudolabel, depending on the training strategy (supervised or self-supervised; see methods for details).

Following transformer encoding, one-dimensional adaptive average pooling was employed to align the output length with the layer’s input. The pooled outputs were further projected to a 256-dimensional embedding, followed by a GELU activation and a final output layer. Masking was performed immediately after the convolutional front-end but prior to positional encoding and transformer processing. The complete model comprised approximately 97.1 million trainable parameters and occupied roughly 404 MB of storage space.

Training proceeded for 40 epochs (i.e., 40 complete passes through the pre-training corpus; chosen to approximately match the number of training steps used in prior SSL work of this type (Hsu et al. 2021)), utilizing a batch size of 64 sequences. Optimization employed the Adam algorithm with a linear learning rate scheduler, gradually increasing from 1e-05 to 5e-04 over the first 15 epochs, then decreasing to 5e-09 over the remaining 25 epochs.

Finally, a second stage of SSL pre-training was performed (as in the original HuBERT formulation; Hsu et al. 2021). Instead of spectral features, k-means labels for the second stage of SSL pre-training were derived from the final transformer layer embeddings of the model trained in the first pre-training stage. This second SSL pre-training stage used the transformer’s native temporal resolution (946 labels per 50.5 minute input sequence, up from 101, a nearly ten-fold increase in resolution), and the difficulty of pre-training was increased by learning a 500-label k-means pseudo-label solution instead of 100 label as in the first stage. The second stage of SSL pre-training started from the model checkpoint produced by the first stage of pre-training (albeit with an updated output layer).

### 2.3. Fine-tuning

To evaluate the health-related information captured by the sleep representations learned via the foundation model’s self-supervised pre-training, we conducted a series of fine-tuning experiments (Fig. 3) to predict health, mood and cognitive variables within the APPLES dataset. An additional set of experiments examined whether SSL pre-training improves data-efficiency by assessing the impact of limiting the size of the downstream dataset (specifically, the number of nights) used to fine-tune models for canonical supervised sleep prediction tasks (ex. AHI, sleep stages), compared to training these models from scratch. In these experiments, the pre-trained SSL model underwent additional training (fine-tuning) for 15 epochs with a modified output layer tailored specifically to each task (similar linear ramp up and down of the learning rate over the first third and final two thirds of epochs, respectively). This process maps the model’s weights and corresponding learned representation to the appropriate, now task-specific, output labels, and further updates the model to optimize task performance. All model weights were updated in this process (i.e., no layers or connections were frozen doing fine-tuning). Regression tasks used a new single-unit output layer (trained with MSE loss, evaluated via Pearson correlation with ground-truth values) whereas classification tasks used a new multi-unit output layer (corresponding to the number of output classes, trained with cross entropy loss, evaluated via prediction accuracy).

**Fig. 3.**
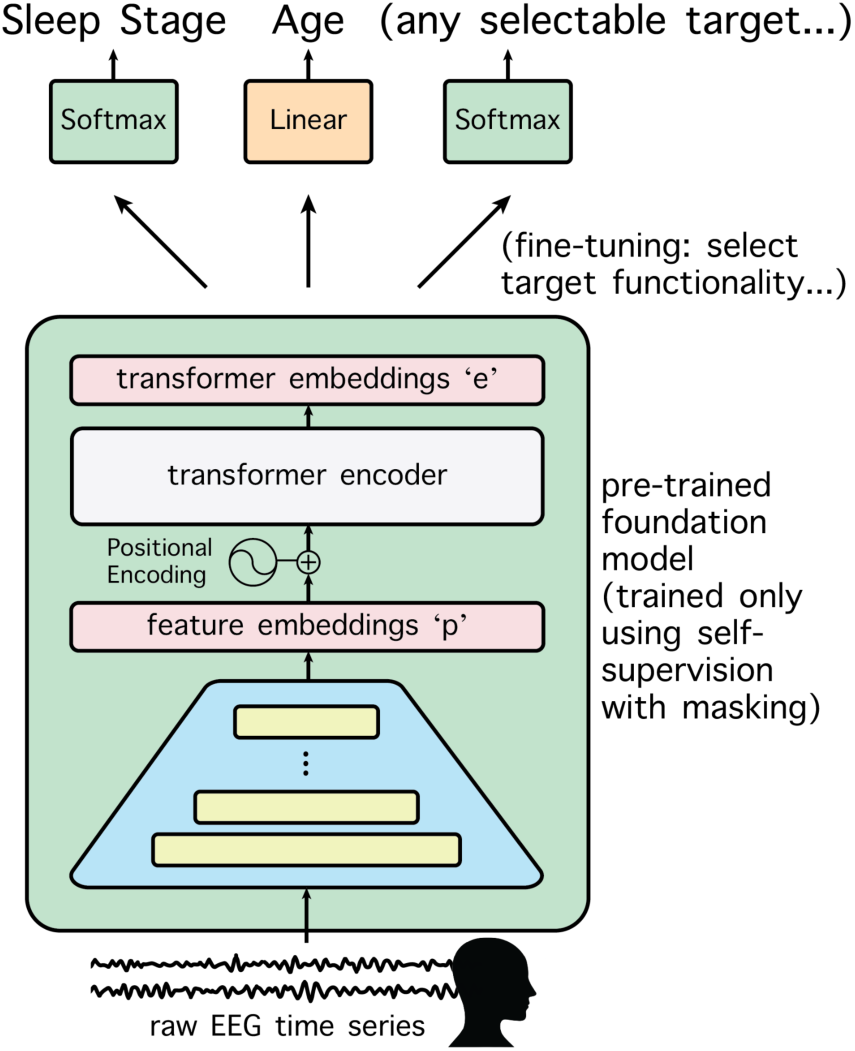
Fine-tuning approach. Models’ information content was probed using downstream predictions tasks, in which pretrained models were further trained (fine-tuned) to predict various health metrics and demographic characteristics.

### 2.4. Baseline Models

To determine whether the SSL-derived, data-driven representations of sleep structure provided richer and more informative content compared to traditional five-stage sleep representations, we pre-trained an identical transformer model explicitly for conventional supervised five-class sleep-stage classification. This model was trained on the same data, utilizing the provided sleep-stage annotations for each polysomnography (PSG) record. This approach allowed us to directly evaluate the richness of the information learned from raw data versus information constrained by traditional sleep-stage labels. Subsequently, we conducted identical fine-tuning experiments with this supervised pre-trained model, enabling a direct comparison of generalizable representations derived from traditional sleep staging against those developed via SSL. The supervised model achieved classification performance comparable to human expert raters, approximately 82% accuracy, consistent with prior reports (Danker-Hopfe et al., 2009; Rosenberg and Van Hout, 2013). The top accuracy for this supervised pre-trained model was 87.7% on the internal validation set, 85.2% on Dreem-H, 85.3% on Dreem-O, 78.1% on HomePAP, and 80.1% on Sleep-EDF datasets.

As a final comparative control, we trained a third set of models identical in architecture to the SSL and supervised pre-trained models but initialized without pre-training. These models were trained from scratch directly on the same prediction tasks utilized during fine-tuning experiments. To ensure competitive performance, these models were trained for an additional 40 epochs (totaling 55 epochs, which equals the total number of training epochs for each of the pretrained models) with the same proportional learning rate schedule, maximizing their opportunity to converge to optimal solutions for each task in the fine-tuning dataset.

To enable statistical quantification of performance differences between models, a 10-fold cross-validation strategy was used to generate distributions of performance values suitable for statistical comparison. Specifically, all models underwent fine-tuning ten times, with each iteration using 80% of the data for training, 10% of the data for validation and the remaining 10% for testing (using the model checkpoint that performed best on the validation data). For the data scaling experiments, models were fine-tuned using 25, 50, 100 or 500 randomly selected nights each. Training was validated using the original validation partition and tested on the APPLES dataset.

## 3. Results

We trained foundation models to autonomously learn high-dimensional representations of structure in sleep data using self-supervised learning (SSL), leveraging a large dataset comprising over 11,000 nights of polysomnographic (PSG) sleep recordings. This SSL task was adapted from methods originally developed for speech recognition. Models were trained on varying combinations of multimodal PSG inputs: EEG alone, EEG and EOG, and a comprehensive set including EEG, EOG, EMG, ECG, and respiratory signals. Here, we primarily discuss models trained exclusively on EEG data, as these demonstrated superior clinical prediction performance, derived the greatest benefit from SSL pre-training, and are most widely applicable to at-home sleep monitoring, which often includes only forehead EEG (Coon, Zerr, et al. 2025; Esfahani et al. 2023; Sikder et al. 2025; Coon and Punjabi 2021). However, similar results were observed for models incorporating additional modalities, as detailed further in the supplementary material.

### 3.1. Health Information is Extensively Encoded in SSL-Derived Sleep Representations

Fine-tuning experiments demonstrated that extensive health-related information could be reliably extracted from sleep EEG data, and that the effectiveness of information extraction varied significantly depending on the deep learning strategy employed (see fine-tuning results and Fig. 4). To assess this, we compared the performance of a self-supervised learning (SSL) approach against control methods predicting clinical outcome metrics and demographic characteristics from the APPLES dataset. We then evaluated the capacity of the SSL-pre-trained foundation model representations to accurately predict each of these outcome measures. Fine-tuning the SSL model enabled above-chance prediction of sleep stage (82.1%, *p* < 0.001), AHI (*r* = 0.54, *p* < 0.001), BMI (*r* = 0.38, *p* < 0.001), age (*r* = 0.68, *p* < 0.001), sex (70.1%, *p* < 0.001), BDI (*r* = 0.04, *p* < 0.01), WASI-IQ (*r* = 0.09, *p* < 0.01), and Pathfinder scores (*r* = 0.09, *p* < 0.001; all values averaged across folds, compared to chance-level performance using Wilcoxon signed rank exact tests).

**Fig. 4.**
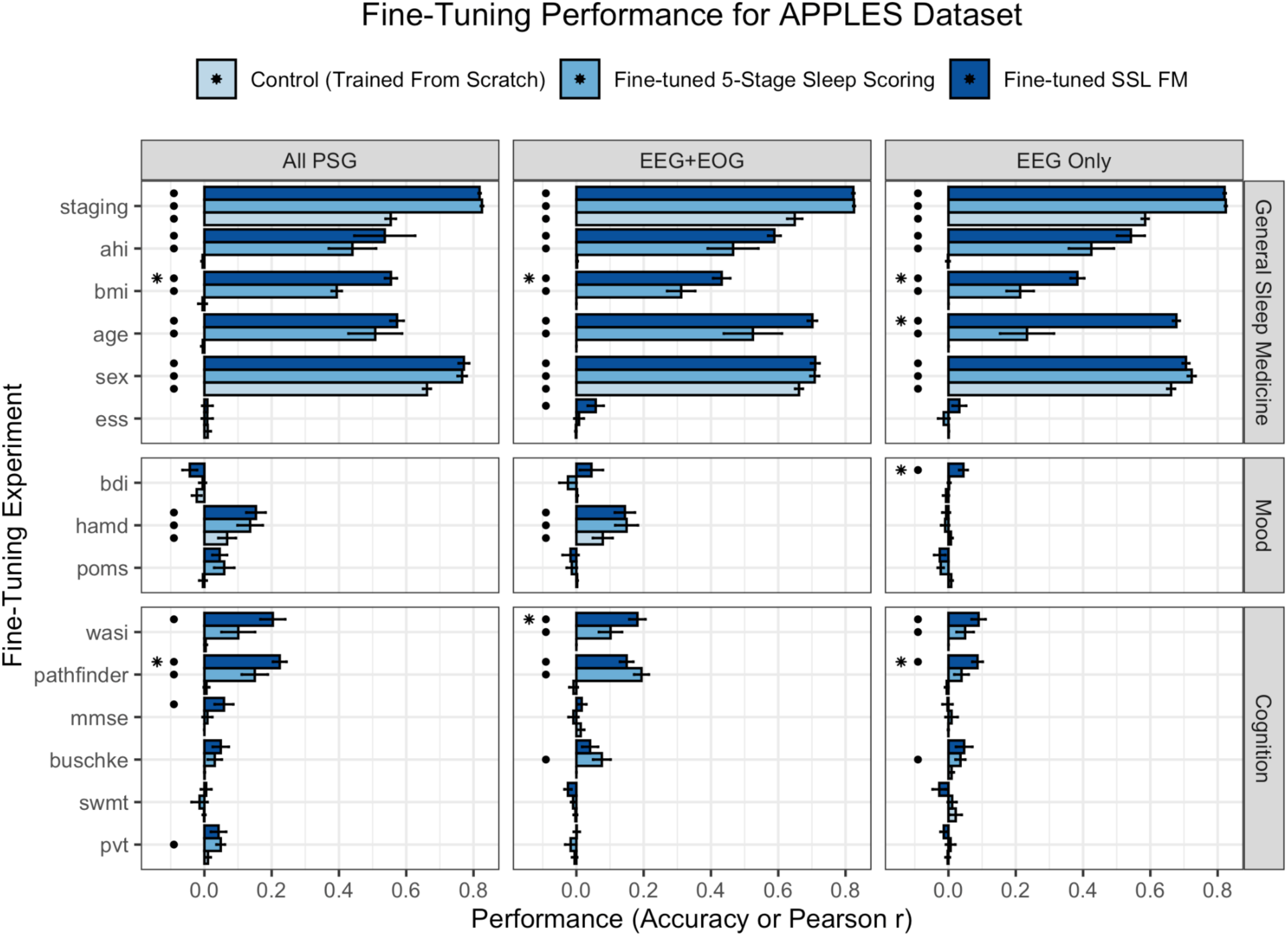
The SSL-trained Foundation Model encodes more health information from sleep data than models trained to internalize traditional sleep stage information. Models pre-trained to identify states through self-supervised learning predict clinical metrics better than models pre-trained from scratch or pre-trained to identify traditional sleep stages. Dots mark fine-tuning experiments that achieve above-chance performance. Asterisks mark tests where, additionally, the SSL model outperformed both baseline models.

### 3.2. Predictive Performance is Not Solely Attributable to Neural Network Architecture

To evaluate whether the predictive capabilities observed in our SSL-derived representation of sleep structure are attributable solely to the neural network architecture itself, we conducted comparative experiments. We reasoned that if the transformer-based models inherently possessed sufficient capacity to directly extract health-related information from sleep EEG signals, then models trained from scratch—without SSL pre-training—to directly predict clinical outcomes (e.g., depression/mood scales, cognitive test scores) should demonstrate comparable predictive performance. Contrary to this, we found that models trained directly on these prediction tasks, without SSL pre-training, exhibited significantly reduced predictive accuracy compared to their SSL pre-trained counterparts. The SSL model out-performed models trained from scratch overall (*V* = 1499, *p* < 0.001) and for predicting specific outcomes including sleep stage classification (*V* = 55, *p* < 0.01), AHI (*V* = 55, *p* < 0.01), BMI (*V* = 55, *p* < 0.01), age (*V* = 55, *p* < 0.01), sex (*V* = 51, *p* < 0.05), BDI (*V* = 45, *p* < 0.01), WASI-IQ (*V* = 53, *p* < 0.01), Pathfinder (*V* = 55, *p* < 0.01; all using paired Wilcoxon signed rank exact tests) (Fig. 4, darkest vs. lightest blue bars).

### 3.3. Predictive Advantage Depends on the Specific Representation of Sleep Structure

Subsequent analyses examined whether the predictive advantage was contingent upon the specific representation of sleep structure—specifically, whether autonomously derived structure via self-supervised learning (SSL) provided superior predictive value compared to traditional sleep staging methods (e.g., conventional five-stage sleep classifications outlined by R&K or AASM guidelines). Direct comparisons demonstrated that the SSL-derived, data-driven sleep representations significantly improved predictive performance relative to traditional staging. This underscores the potential of SSL methods to capture nuanced physiological dynamics beyond those represented by standard sleep scoring frameworks. (Fig. 4, comparison between the two darkest blue bars). The SSL model outperformed the model pre-trained for traditional supervised sleep stage classification overall (*V* = 3383, *p* < 0.001) and for predicting specific outcomes including BMI (*V* = 55, *p* < 0.01), age (*V* = 54, *p* < 0.01), BDI (*V* = 48, *p* < 0.05), Pathfinder (*V* = 47, *p* < 0.05). No significant difference in performance was found between the SSL and supervised sleep stage models for AHI (*V* = 44, *p* = 0.11), sex (*V* = 11, *p* = 0.11), WASI-IQ (*V* = 41, *p* = 0.19); all using paired Wilcoxon signed rank exact tests). Finally, the SSL model was marginally outperformed (82.4% and 82.1%) by the supervised sleep stage classification model for sleep stage classification (*V* = 6, *p* < 0.05).

### 3.4. SSL-Derived Sleep Structure Exhibits Greater Complexity Than Traditional Stages

To further investigate the complexity of the sleep structure identified by SSL pretraining, we employed a Bayesian Information Criterion (BIC)-based analysis. BIC scores were calculated across a range of cluster solutions, from 0 to 50 states based on k-means clustering of activations from the SSL model’s final transformer layer for each PSG record, providing a metric that balances model fit against model complexity. Minimum BIC values indicate an optimal number of states necessary to capture the underlying structure. Individual recordings were analyzed separately, resulting in an optimal average fit of 10-11 states (Median = 10, IQR = 3 for the HomePAP data; Median = 11, IQR = 3 for the APPLES data; Figure 5A). As a control analysis we ran the same BIC cluster-sweep analysis on the supervised model pre-trained for traditional five-stage sleep classification. This yielded an optimal number of five clusters (Median = 5, IQR = 1 for the HomePAP data; Median = 5, IQR = 1 for the APPLES data; Figure 5A), reflecting the expected solution aligned with the five stages the model was trained to identify. The distribution of BIC-optimal states derived from the SSL model was significantly higher than for the supervised baseline models (*W* = 7132.5, *p* < 0.001 for the APPLES dataset, *W* = 594.5, *p* < 0.001 for the HomePAP dataset, both Wilcoxon rank sum tests), suggesting the SSL model had learned a more complex representation of sleep physiology than models trained to predict sleep stages via traditional methods.

**Fig. 5.**
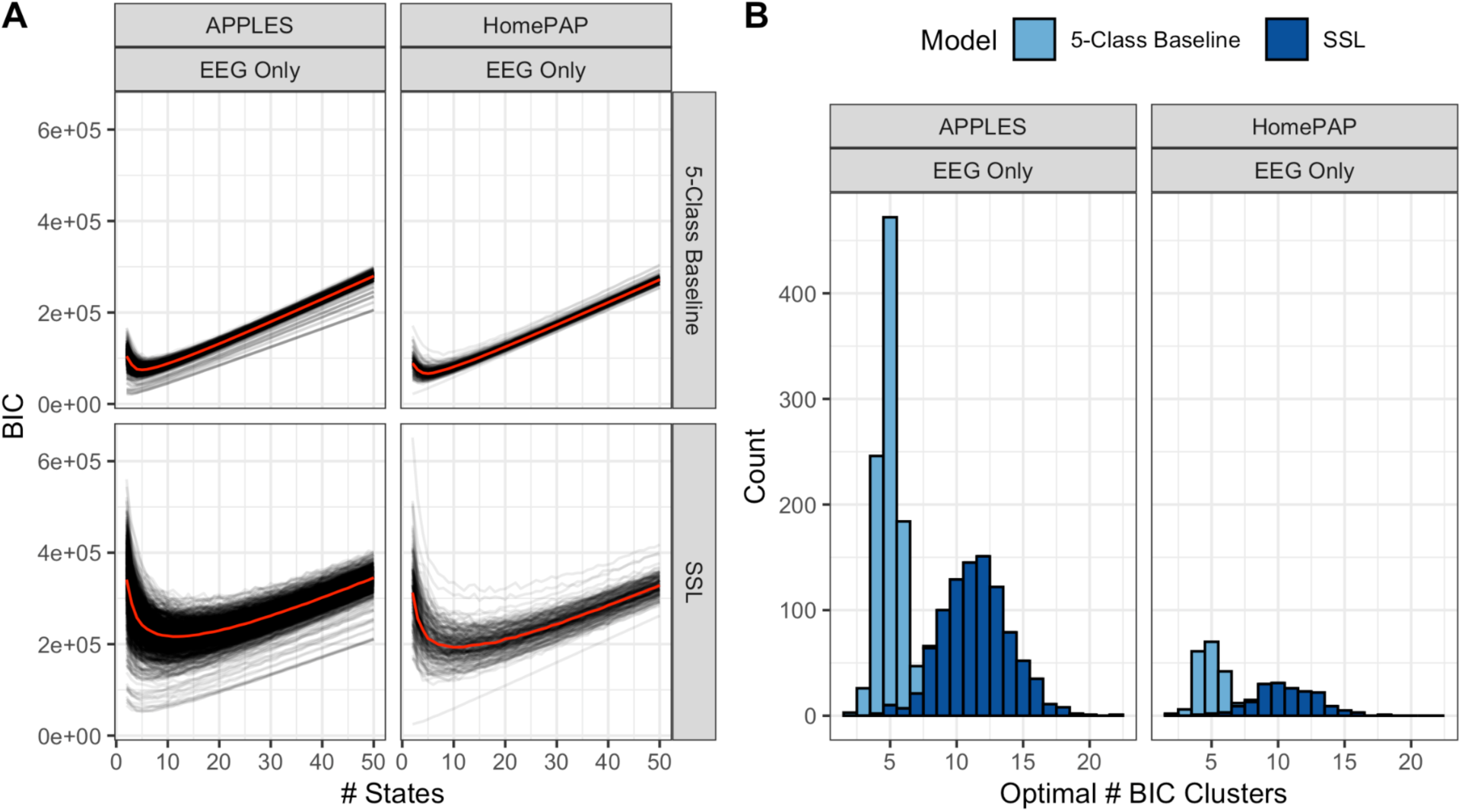
Foundation Models trained using SSL learn more complex representations of sleep structure than models trained on the conventional supervised sleep stage labeling task. A) BIC values as a function of the number of k-means clusters used to describe the model embedding space projections of the data in the APPLES and HomePAP datasets. Black lines show individual PSG records. Red lines show median across records in each dataset. B) Distribution of the optimal (minimum) BIC values obtained for each PSG record in the APPLES and HomePAP data colored by model. The optimal cluster number is significantly higher for the SSL model’s representation of sleep data (10-11 clusters (see Section 3.4 for details) compared to the (expected) 5 that emerge from conventional sleep staging models).

### 3.5 SSL Pre-Trained Models Allow Efficient Use of Labeled Data

In addition to autonomously identifying state structure in unlabeled data, foundation models frequently exhibit superior performance on downstream tasks due to their extensive, application-agnostic pretraining. Minimizing reliance on labeled data can significantly expedite research and facilitate the development of sleep assessments. To examine potential gains in fine-tuning efficiency provided by our SSL-pretrained model, we conducted an additional series of experiments that utilized smaller datasets, specifically focusing on canonical supervised sleep prediction tasks. Our results indicate that SSL pretraining markedly improves model performance with limited amounts of labeled data compared to both models trained from scratch and those pretrained on supervised sleep-stage classification (Fig. 6). The SSL-pretrained model achieved above-chance performance with data from as few as 25 records in several cases (AHI and sleep-stage classification both V = 15, p < 0.05), and in all evaluated tasks when using 50 records (age V = 15, p < 0.05). With 100 records, the SSL-pretrained EEG model consistently outperformed both baseline models (all W > 24, p < 0.05). Notably, the model fine-tuned for sleep-stage classification achieved high accuracy even with data from only 25 PSG records, surpassing performance of a model trained from scratch in all tested scenarios (W = 25, p < 0.01). The supervised sleep-stage classification pretrained model was not included in this fine-tuning comparison because it was already pretrained on this task.

**Fig. 6.**
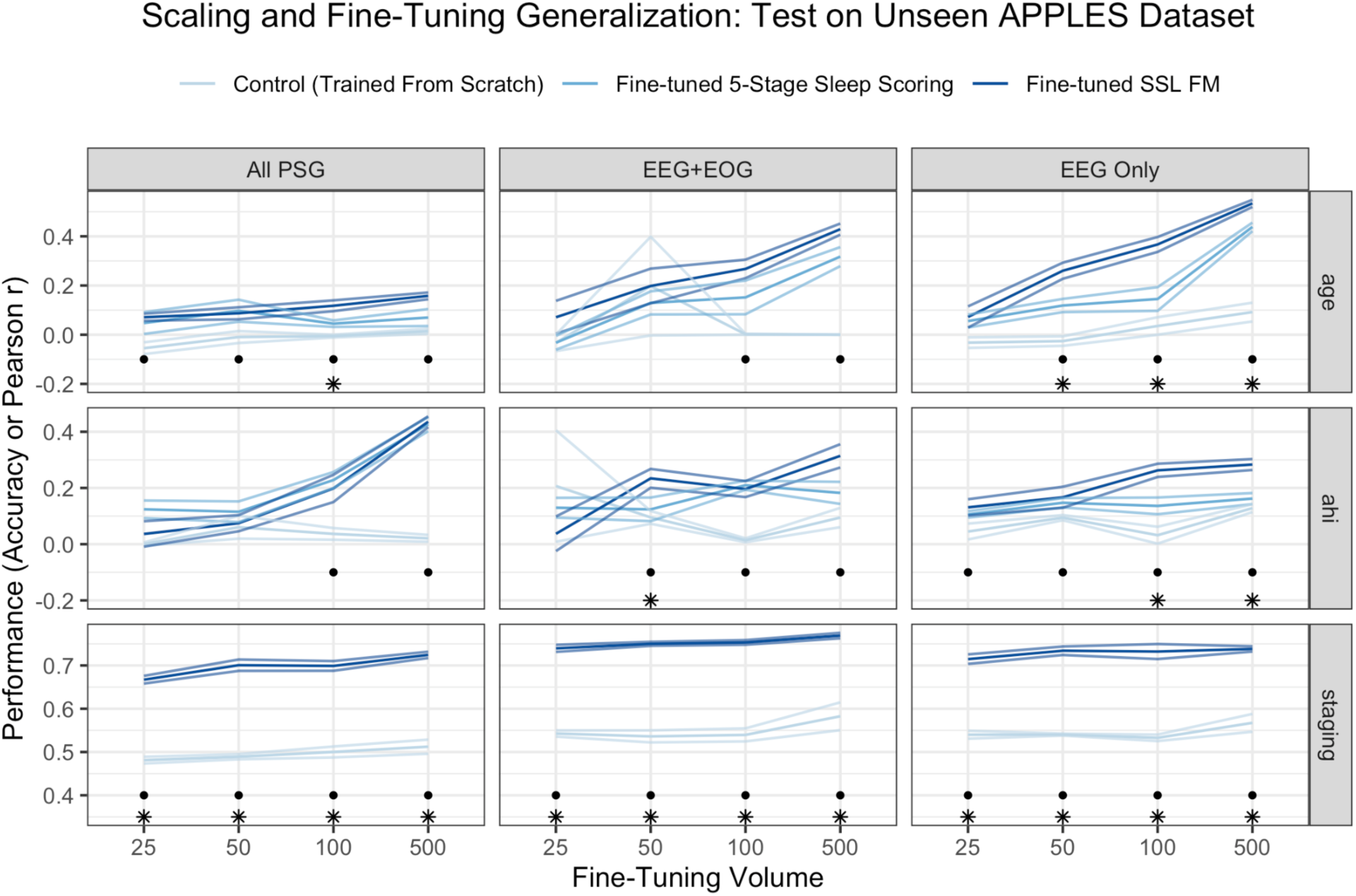
Models pre-trained to learn to identify states through self-supervised learning can be fine-tuned to predict canonical sleep tasks on downstream tasks with limited labeled data. Dots mark fine-tuning experiments where the SSL model achieved above-chance performance. Asterisks mark tests where the SSL model performed above chance and outperformed both baseline models.

### 3.6 SSL Pre-Trained Models Autonomously Capture the Gross Structure of Sleep

The strong performance of the SSL model on downstream sleep-stage classification tasks, even when fine-tuned on as few as 25 overnights, indicates that its autonomously learned representation of sleep inherently captures the overarching physiological structure that underpins traditional sleep staging systems. To further substantiate this, we conducted a high-level comparative analysis using two benchmark datasets (Dreem and Sleep-EDF). Data from these corpora were projected through both the SSL-pretrained model and the supervised sleep-scoring baseline model prior to fine-tuning. The resulting transformer model embeddings for each epoch were visualized using a two-dimensional t-Distributed Stochastic Neighbor Embedding (t-SNE; van der Maaten & Hinton, 2008). Figure 7 presents these visualizations side-by-side, organized according to their respective ground-truth sleep stage annotations. The visualization reveals notable similarities in the global sleep structure learned independently by both models. This clearly demonstrates that the SSL model autonomously captures the primary physiological distinctions underlying traditional sleep stages, without explicit reliance on sleep-stage labels during pre-training. This, in turn, confirms that the SSL-derived sleep states indeed align closely at a gross level with established sleep stages, while also highlighting the additional, nuanced structure identified by the SSL methodology.

**Fig. 7.**
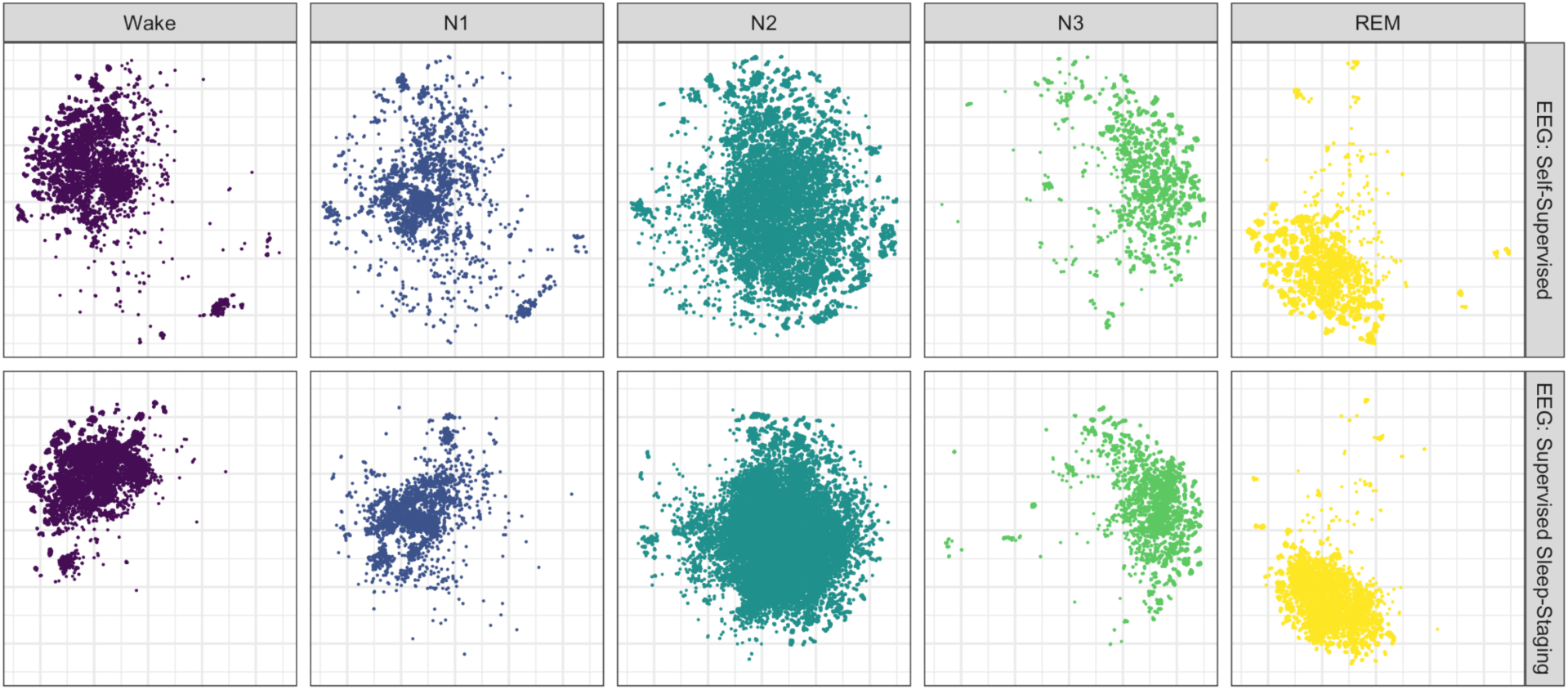
t-SNE visualization of EEG (Dreem data) reveals that models pre-trained using SSL methods learn a global structure of sleep similar to that learned by models trained to predict sleep stages using supervised methods, despite the absence of any sleep-stage labels in SSL pre-training. A similar pattern of results was obtained from this analysis when run on the Sleep-EDF dataset.

## 4. Discussion

This study demonstrates the significant potential of leveraging self-supervised learning (SSL) techniques, specifically using Foundation Models (FMs), to derive more nuanced, physiologically meaningful representations of sleep structure from EEG data. Traditional sleep staging methods, while still crucial to the practice of modern sleep medicine, remain limited by inherent subjectivity, low temporal resolution, and coarse categorization, neglecting subtle and continuous variations within sleep stages. The SSL approaches utilized here address many of these limitations by autonomously identifying more complex structure from sleep data, which provides enhanced predictive capabilities for multiple clinical and cognitive health metrics.

A growing body of prior work has begun to explore self-supervised learning (SSL) methods to train foundation models specifically for sleep data. These have predominantly concentrated on replicating traditional sleep stages (Fox et al., 2025) or involved limited baseline comparisons (Thapa et al., 2024). Consequently, the full potential of SSL-trained foundation models relative to traditional supervised sleep-stage classifiers has remained underexplored. To our knowledge, this study provides the first thorough evaluation comparing an SSL-based sleep foundation model directly against a strong supervised baseline model fine-tuned explicitly for traditional five-class sleep stage classification across various downstream prediction tasks. Additionally, this research represents the most extensive exploration thus far into fine-tuning supervised sleep-stage classification models for tasks extending beyond sleep staging itself. We demonstrate that fine-tuned supervised sleep-stage classifiers effectively handle several downstream tasks, often outperforming models trained directly from scratch. Specifically, supervised models showed above-chance performance on multiple tasks, including age prediction, sex classification, and HAM-D prediction. Our analyses nevertheless demonstrate that self-supervised pre-training provides advantages over fine-tuned sleep-stage classifiers, as it facilitates the learning of flexible, generalizable representations that not only capture the broad physiological structure of sleep but also incorporate additional clinically relevant features, significantly enhancing performance on various downstream tasks. Taken together, these results not only reinforce prior findings that SSL models are effective for downstream sleep-stage classification (Fox et al., 2025) and related tasks (Thapa et al., 2024), but also expand upon prior work by assessing performance in cognitive and mood-related areas, supported by robust baseline comparisons with traditional supervised methods.

A key finding of this study was the substantial superiority of SSL-derived representations of sleep when predicting measures of health (AHI, BMI), mood (BDI), and cognition (WASI, Pathfinder, Buschke), compared to representations derived from standard supervised five-stage sleep classifier models, or to task-specific prediction models initialized from scratch without pre-training. These comparisons suggest that Foundation Models autonomously learn information about health and brain function from sleep data that is overlooked by traditional sleep analyses. The implications of this extend beyond sleep science, suggesting that there may still be substantial untapped value even in exhaustively mined datasets, when exposed to Foundation Model training techniques.

A critical aspect of this finding emerged from control analyses demonstrating that the enhanced predictive performance of SSL models cannot be explained merely by the architecture of the neural network or extended training alone. Models trained extensively from scratch failed to replicate the predictive accuracy of those pretrained via SSL, strongly indicating that the predictive strengths of the model are rooted in the autonomously internalized, SSL-derived latent representations of sleep. Put simply, the key information is embedded uniquely in the data-driven SSL representation itself—not in just any representation of sleep—as evidenced by the fact that pre-training on standard 5-class AASM sleep stages did not confer comparable predictive benefits.

In addition to this central result, secondary analyses revealed other noteworthy benefits of SSL pretraining. SSL models significantly outperformed traditional methods in predicting Apnea-Hypopnea Index (AHI, particularly when data is limited), age, and body mass index (BMI) using EEG alone. Traditionally, estimating AHI requires cumbersome equipment typically available only in clinical settings; thus, the model’s capability to achieve accurate estimates using only EEG data highlights EEG’s potential as an affordable, comfortable, and effective tool for at-home diagnostic screening.

Furthermore, the SSL model’s ability to accurately predict biological age from EEG underscores its utility in developing novel biomarkers, such as measuring the brain-age gap (Cole and Franke 2017; D. Zhang et al. 2024; Brink-Kjaer et al. 2022; Banville et al. 2024), which may offer critical insights into brain health, cognitive function, and aging. Given the growing importance of age-related neural biomarkers in predicting cognitive decline and the expanding availability of at-home EEG wearables, these findings suggest practical, scalable opportunities for widespread clinical deployment.

In summary, this study demonstrates the significant potential of self-supervised learning (SSL) techniques, specifically through Foundation Models (FMs), to autonomously derive nuanced, physiologically meaningful representations of sleep structure from PSG data. While traditional sleep staging remains invaluable, it is also inherently limited by subjective interpretations, low temporal resolution, and coarse categorization, neglecting subtle variations within sleep states. SSL-based Foundation Models, in contrast, independently uncover complex sleep structure that supports enhanced predictive capacities on diverse health, cognitive, and mood-related outcomes. Crucially, the predictive power of these models is specifically attributable to the SSL-derived latent representations, rather than merely to network complexity or extensive training alone. These findings underscore that autonomously learned SSL representations uniquely capture clinically and cognitively relevant information overlooked by traditional sleep analyses, highlighting substantial untapped potential within sleep data when explored through hypothesis-free, data-driven SSL approaches.

## Data Availability

This study produced no new data. All data analyzed were publicly available at the National Sleep Research Resource (NSRR) at sleepdata.org; the Dreem Open Datasets (https://dreem-dodo-dodh.s3.eu-west-1.amazonaws.com/index.html); and Physionet's Sleep-EDF dataset at https://www.physionet.org/content/sleep-edfx/1.0.0/

https://sleepdata.org

https://dreem-dodo-dodh.s3.eu-west-1.amazonaws.com/index.html

https://www.physionet.org/content/sleep-edfx/1.0.0/

